# Accelerated Biological Aging in Familial Pulmonary Fibrosis

**DOI:** 10.1101/2024.10.10.24315235

**Authors:** Ana P.M. Serezani, Daphne B. Mitchell, Joy D. Cogan, M. Merced U. Malabanan, Cheryl R. Markin, Errine T. Garnett, Katrina Douglas, Tisra H. Fadely, Jonathan A. Kropski, Margaret L. Salisbury, Timothy S. Blackwell

## Abstract

Since Familial Pulmonary Fibrosis (FPF) manifests in older adults and telomere attrition is common in FPF and sporadic Idiopathic Pulmonary Fibrosis (IPF), we postulated that accelerated aging, as determined by epigenetic clock (DNA methylation) measurements, could occur in FPF. We measured DNAge from blood of patients with FPF and a group of first-degree relatives of FPF patients without disease (termed “at-risk” for FPF) with or without genetic rare variants (RVs) in telomerase pathway genes. We observed accelerated epigenetic aging with increased DNAge compared to chronological age in individuals at-risk for FPF and FPF patients compared to healthy controls. We found that increased DNAge manifests independently of the presence of RVs in telomerase pathway genes or telomere length in peripheral blood cells. These findings suggest that increased DNAge could be an independent risk factor for the development of FPF.

## Introduction

Aging is a risk factor for Idiopathic Pulmonary Fibrosis (IPF) and its familial form, Familial Pulmonary Fibrosis (FPF) (1, 2). Heterozygous loss-of-function genetic rare variants (RV) in telomere maintenance genes, which can lead to telomere shortening, are the most common genetic mechanism driving FPF. In addition, short telomeres (below the 10th percentile) in peripheral blood cells and epithelial cells in the lung are present in forty percent of patients with FPF and twenty-five percent of IPF (3, 4). We recently reported that asymptomatic first-degree relatives of patients with FPF (termed “at-risk relatives”) have a faster annual decline in telomere length than expected for the population (5), suggesting that age-mediated processes could predate the development of clinical disease.

Aging is the natural and progressive change in a variety of cellular functions, causing increased vulnerability to disease and limiting lifespan (6). Accumulation of epigenetic changes in chromatin is a hallmark of aging, which can influence gene expression in ways that impact cellular functions (7). Changes to DNA methylation at specific CpG sites have been proposed as a reliable estimation of the biological age of cells and tissues, termed Epigenetic Clock (8).

Biological age-predictive models have been developed based on methylation patterns across loci in different tissues from healthy subjects (8). While most individuals have a concordant biological and chronological age, some exhibit accelerated epigenetic aging, particularly during chronic diseases (9-12). In this study, we measured the epigenetic age in patients with FPF and their asymptomatic relatives.

## Results

These studies included 56 patients with FPF, 54 asymptomatic relatives, and 23 controls without a personal or family history of pulmonary fibrosis. All included members of FPF families had genetic sequencing to identify pathogenic RVs in telomerase genes (*TERT, TERC, RTEL1, PARN*) and peripheral blood cells telomere length. Participant characteristics are in **Fig 1A**. We observed a greater discrepancy between the epigenetic and chronological ages (termed ΔDNAge, calculated as epigenetic age minus chronological age) in at-risk relatives (6.6±5 years) and subjects with FPF (4.4±6 years) compared to control subjects (0.3±4 years), indicating accelerated epigenetic aging in FPF kindreds (**Fig 1B**). Using a linear regression analysis, we found little correlation (R^2^=0.019, p=0.14) between ΔDNAge and telomere length (percentile corrected for chronological age) in peripheral blood cells among members of families with FPF (**Fig 1C**); however, FPF patients with very short telomeres (at the 1^st^ percentile for age) had greater ΔDNAge compared to those with longer telomeres (**Fig 1D**). In addition, there was no difference in ΔDNAge among FPF family members with or without a pathogenic telomere-related gene RV (**Fig 1E)**. Among at-risk relatives, there was no difference in ΔDNAge between those with or without evidence of subclinical FPF (i.e., interstitial lung abnormalities) on CT scan (**Fig 1F**).

**Figure 1.**
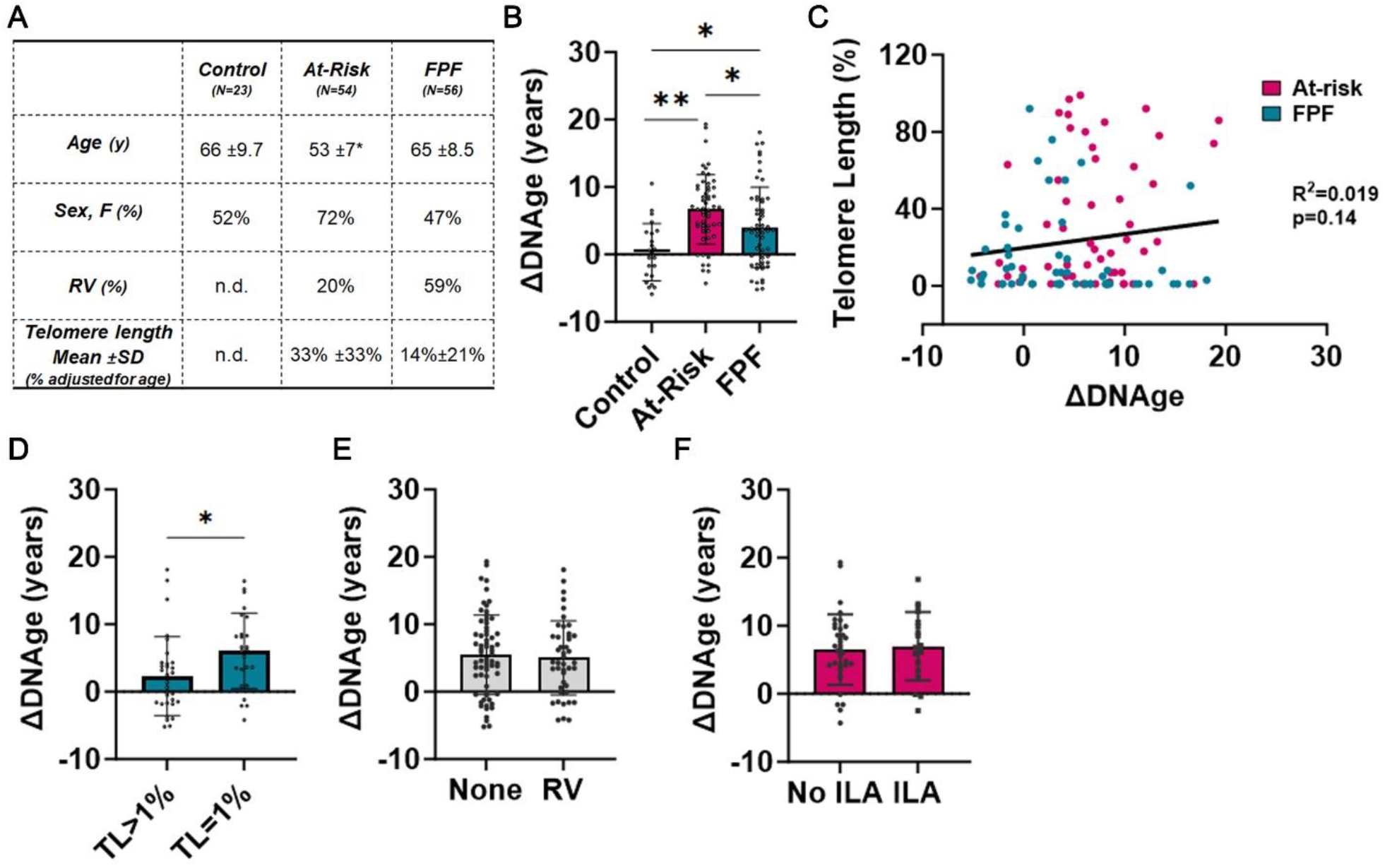
Epigenetic analysis of subjects with FPF and at-risk relatives indicate accelerated biological aging. **(A)** Characteristics of participants selected for DNA-associated aging analysis (n.d. = not done). (**B)** ΔDNAge (difference between Epigenetic Age and chronological age) determined from DNA isolated from whole blood of healthy controls, at-risk subjects in FPF kindreds and FPF patients. **(C)** Linear regression analysis of ΔDNAge and telomere length (TL) from at-risk family members (magenta) and FPF (green) patients. **(D)** ΔDNAge of FPF subjects with TL=1% or TL>1%. **(E)** ΔDNAge of participants with and without heterozygous rare variants (RV) in telomerase pathway genes. **(F)** ΔDNAge of at-risk family members with and without interstitial lung abnormalities (ILA) on CT scan. Mean ± SD are shown. *p < 0.05, **p < 0.0001 [one-way ANOVA]. Chi-square was used for categorical variables.

## Discussion

In this study, we used Epigenetic Clock measurements to examine the role of biological (epigenetic) aging in familial pulmonary fibrosis. Since telomerase pathway function and telomere length have been associated with aging in pulmonary fibrosis, we compared epigenic aging with the presence of telomerase pathway gene RVs and peripheral blood cell telomere length and showed that exaggerated epigenetic aging is a feature of FPF. There was little correlation between ΔDNAge and telomere length or the presence of telomerase pathway RVs. In addition, exaggerated epigenetic ageing was present in at-risk relatives. Interestingly, relatives with evidence of subclinical FPF on the enrollment screening examination had no difference in ΔDNAge than those without subclinical disease, suggesting that accelerated epigenetic age may be present in at-risk family members prior the onset of pathologic lung changes detectable by CT imaging. While this study provides valuable insights about biological aging in families with FPF, limitations include the size of the cohort analyzed and the absence of a longitudinal analysis, which impair a complete evaluation of biological aging progression in FPF. We did not evaluate the impact of epigenetic aging on disease severity or survival time in patients with FPF, nor did we examine the influence of aging on cellular functions. However, our study has shown that FPF might be influenced by biological aging processes other than telomere shortening. Unraveling the causes of aging could significantly contribute to defining early disease mechanisms and prevention strategies.

## Materials and Methods

### Familial Pulmonary Fibrosis Registry Methods

Participants in these analyses were selected from the Familial Pulmonary Fibrosis (FPF) Registry at Vanderbilt University Medical Center, which enrolls bloodline members of families in which 2 or more members have an idiopathic interstitial pneumonia, including at least one with idiopathic pulmonary fibrosis (IPF). IPF is diagnosed according to American Thoracic Society/European Respiratory Society consensus criteria (13). Asymptomatic first-degree relatives (termed “relatives”) of patients with FPF are eligible to participate in an ongoing observational study that conducts serial screening examinations (high-resolution chest CT [HRCT] and pulmonary function test [PFT]) to identify subclinical FPF (14). Subclinical FPF is defined by the presence of interstitial lung abnormalities (ILAs) on HRCT (15). The FPF registry also enrolls control participants without a personal or family history of pulmonary fibrosis. These studies were approved by the Vanderbilt University Institutional Review Board (IRB# 020343, 080780).

### Telomere length

The mean telomere terminal restriction fragment length (MTL) was measured in DNA extracted from whole blood via southern -blot and percentile for age was calculated using a healthy control population as we have previously described (15).

### Biological Age using Epigenetic clock

Genomic DNA was isolated from whole blood samples using Gentra puregene kit (Qiagen). DNA samples were submitted to Zymo Research to measure epigenetic age. Bisulfite conversion of DNA was performed according to the standard protocol (EZ DNA Methylation-Lightning™ Kit) to differentiate unmethylated from methylated cytosines residues. High throughput methylation analysis followed by DNA methylation age analysis were performed using the Simplified Whole-panel Amplification Reaction Method (16). Data was analyzed using elastic net regression of DNA methylation levels (Delta DNA methylation) according to Zymo Research’s proprietary DNAge® predictor. DNA methylation levels of 353 CpG sites (9) were used to calculate DNAge (adjusted biological age). Results were expressed as ΔDNAge, where ΔDNAge = DNAge – chronological age at the time of sampling.

### Statistics

All data were presented as mean ± standard deviation (SD). Comparisons between groups were analyzed by unpaired two-tailed Student’s t test or two-way ANOVA. Simple linear regression analysis with 95% interval was performed for correlation between telomere length and ΔDNAge®. Statistical analysis was performed using Prism 10 (GraphPad).

## Data Availability

All data produced in the present study are available upon reasonable request to the authors

## Acknowledgments

We would like to thank FPF families for participating in this study. We would like to thank Dr. Amy Major, Dr. Digna Velez Edwards, Dr. Andreanna Holowatyj, Dr. Jacklyn Hellwege, Dr. Kelsie Full, Dr. Elizabeth Jasper, Dr. Megan H. Shuey, and Dr. Lauren Wareham for insightful comments.

## Notes

### Competing Interest Statement

The authors have declared no competing interest.

### Funding Statement

5K12HD043483-12
P01HL172729

### Author Declarations

These studies were approved by the Vanderbilt University Institutional Review Board (IRB# 020343, 080780).

